# Generative Artificial Intelligence in Psychotherapy Practice: A Global Online Survey of Mental Health Professionals’ Adoption

**DOI:** 10.64898/2026.06.18.26355962

**Authors:** Charlotte Blease, Josefin Hagström, Jens Gaab, Alexis Carey, Francesco Cipriani, Colin Gorman, Antje Frey Nascimento, Amanda Fitzgerald, Lena Holtz, Julian Schwarz, Gillian Strudwick, Maria Tibbs, John Torous, Jeffrey H. D. Cornelius-White

## Abstract

**Background:** Generative artificial intelligence (GenAI) tools, including large language model (LLM)-based platforms such as ChatGPT, Google Gemini, and Microsoft Copilot, are being adopted across healthcare settings with increasing speed. Despite the increasing popularity of GenAI, empirical data on the extent and nature of adoption by mental health clinicians in routine psychotherapy practice globally remain scarce.

**Objective:** This study aimed to characterize current use patterns of GenAI tools among a global sample of practicing mental health professionals, including prevalence of use, specific tools employed, clinical and administrative purposes served, perceived effect on workload, and the institutional context shaping adoption (e.g., encouragement, prohibition, and training).

**Methods:** We administered a cross-sectional online survey to a global convenience sample of licensed mental health professionals who provide psychotherapy as part of the scope of their practice (i.e., psychotherapists, psychologists, counsellors, nurses, and psychiatrists). Participants were recruited via professional networks, purposely avoiding the use of social media platforms. Within the survey, we captured GenAI use behaviors in psychotherapy contexts, and demographic and professional background data. Descriptive statistics were analyzed for all variables. Multivariate logistic regression was used to examine demographic and professional predictors of GenAI use.

**Results:** A total of 766 mental health professionals who provide psychotherapy from 30 countries completed the survey. Of these, 54.6% (n=418) reported having purposely used at least one GenAI tool in psychotherapy clinical practice. ChatGPT was the most frequently used tool (354/418, 84.7%). The most commonly reported clinical purpose was assisting with treatment planning (175/418, 41.9%), followed by managing administrative tasks (173/418, 41.4%) and generating psychoeducational materials for clients (166/418, 39.7%). 82.8% of AI users reported that these tools reduced their overall work burden. Only 18.1% (139/766) of respondents reported institutional encouragement to use AI tools, while 81.1% (621/766) reported not having received any professional training on AI use. Predictors of AI adoption included younger age and rural practice setting.

**Conclusions:** In this global convenience sample survey, GenAI use among mental health professionals in psychotherapy settings is widespread, concentrated in a wide variety of clinical and administrative tasks. Formal training and institutional guidance substantially lag behind current adoption patterns. These findings highlight an urgent need for evidence-based competency frameworks, regulatory clarity, and professional education to support safe and ethically informed integration of AI into clinical mental health practice.

## Introduction

The emergence of generative artificial intelligence (‘GenAI’), defined here as AI systems capable of producing text, summaries, or other content in response to natural language prompts, has precipitated rapid and largely unregulated adoption across healthcare professions [1–4]. Platforms such as OpenAI’s ChatGPT, Google’s Gemini, Claude AI, and Microsoft Copilot are now accessible to clinicians and patients alike, with minimal technical barriers to entry [5]. In parallel, ambient scribe tools that incorporate GenAI tools, and which passively capture and process clinical conversations in real time, have begun to enter clinical workflows, raising distinct questions about consent, accuracy, and relational dynamics [6].

Mental health practice presents a particular case study in GenAI adoption. LLMs are experienced by some users as a relationship rather than a tool: emerging empirical work documents the formation of parasocial bonds [7–9]. Indeed, unlike many clinical approaches, psychotherapy is fundamentally relational: positive regard, therapeutic alliance, empathic attunement, and client-centered responsiveness are not incidental features but central mechanisms of effective treatment [10,11]. In addition, practitioners working in psychotherapy contexts also face significant documentation and administrative burdens that AI tools may be able to meaningfully alleviate [8].

Prior surveys of AI attitudes and behaviors among health professionals have predominantly sampled physicians in high-income countries [12–14]. In the era of GenAI, surveys of mental health professionals specifically are nascent. However, in North America this is beginning to change. In 2025, 56% of American Psychological Association psychologists reported using these tools in practice [15]. Another qualitative interview study of 18 US psychotherapists found that trust in GenAI was conditional and task-dependent, sustained for low-stakes administrative work like documentation and brainstorming, but withdrawn when AI threatened clinician control, and clinical judgment [16]. In another recent US study, 47% of American Psychiatric Association members reported using these tools [4]. Despite this, European, and global data are largely absent, and no study to date has examined the full expanse of clinical AI use - tools employed, purposes served, perceived workload impact, institutional context, and training exposure - across a broadly international sample.

This study addresses that gap. Drawing on a global convenience sample of practicing mental health professionals, we characterize the landscape of GenAI adoption in psychotherapy clinical practice in the first quarter of 2026. Our findings are intended to inform policymakers, professional bodies, and training programs seeking to develop evidence-based responses to GenAI integration in mental health services.

### Research Objectives

1. To determine the prevalence of purposeful GenAI use among global mental health professionals.
2. To identify the specific GenAI tools in use and the clinical and administrative purposes they serve.
3. To assess whether clinicians perceive GenAI tools as reducing their work burden.
4. To characterize the institutional context of GenAI use, including employer encouragement, prohibition, and formal training provision.
5. To examine demographic and professional predictors of GenAI adoption.

## Methods

### Study Design

This is a pre-registered cross-sectional mixed methods survey study conducted between January and March 2026. The survey was administered online via LimeSurvey (LimeSurvey GmbH, Hamburg, Germany). This paper reports findings from Sections A (AI use behaviors) and D (demographics) of a larger questionnaire (for the full survey instrument see Multimedia Appendix 1). This study was conducted and reported in accordance with the CHERRIES checklist for online health surveys [13] (see Multimedia Appendix 2).

### Ethics Approval

Ethical approval was obtained from the Institutional Review Board of the Faculty of Psychology, University of Basel (IRB number: 017-25-1) and Missouri State University’s Institutional Review Board (IRB-FY2025-325). All participants provided informed electronic consent prior to survey commencement. Participation was voluntary, anonymous, and unpaid. Personally identifying information—including name, contact details, and IP address—was not collected. LimeSurvey is a secure online platform that employs encryption and anonymization to prevent survey responses from being linked to individual participants. All identifying information, including email addresses, was removed prior to the dataset being shared with the research team. The platform operates in compliance with the European Union General Data Protection Regulation (GDPR).

### Participants and Eligibility

Eligible participants were trained mental health professionals—including psychotherapists, psychologists (clinical and counseling), counselors, psychiatrists incorporating psychotherapy, social workers providing psychotherapy, nurses providing psychotherapy, and other self-identified practitioners of psychotherapy—who were currently practicing or had practiced within the prior 12 months. Participants were required to be aged 18 years or older and to provide informed consent electronically. There were no country-of-practice restrictions.

### Recruitment

Participants were recruited via a global convenience sampling strategy between 13 January 2026 and 30 March 2026 (approximately 10 weeks). The survey link was disseminated directly and through snowball-sampling requests to professional contacts via e-mail. Recruitment channels included professional associations and networks (psychology, psychiatry, counselling, and social work bodies), academic and clinical mailing lists predominantly in Austria, Canada, Ireland, Germany, Switzerland, the UK, and the USA. We deliberately avoided use of all social media platforms to administer the survey to avoid the risk of bot contamination. Minimum target sample size was calculated a priori using standard power analysis for proportions (anticipated AI use prevalence = 30%; margin of error = 5%; 95% CI; N ≈ 323), with a target of ≥500 completed responses to enable subgroup analyses.

### Survey Instrument

The survey was developed iteratively by the lead members of the research team (CB, J C-W, and JG) with expertise in digital mental health, clinical psychology and psychotherapy, and survey methodology. Items were piloted with a convenience sample of three practicing clinicians from the US, UK, and Sweden before finalization. The survey was timed to be approximately 5 to 7 minutes to complete and included four sections - Section A encompassed GenAI adoption (see below), Section B included items focusing on participants’ opinions about GenAI in psychotherapy, Section C included an open comment question, and Section D focused on demographic questions. All closed-ended items were mandatory, while one open-ended free-text question was optional. For this paper, the following sections of the survey are reported (see Table 1, also Multimedia Supplementary File 1).

**Table 1:**
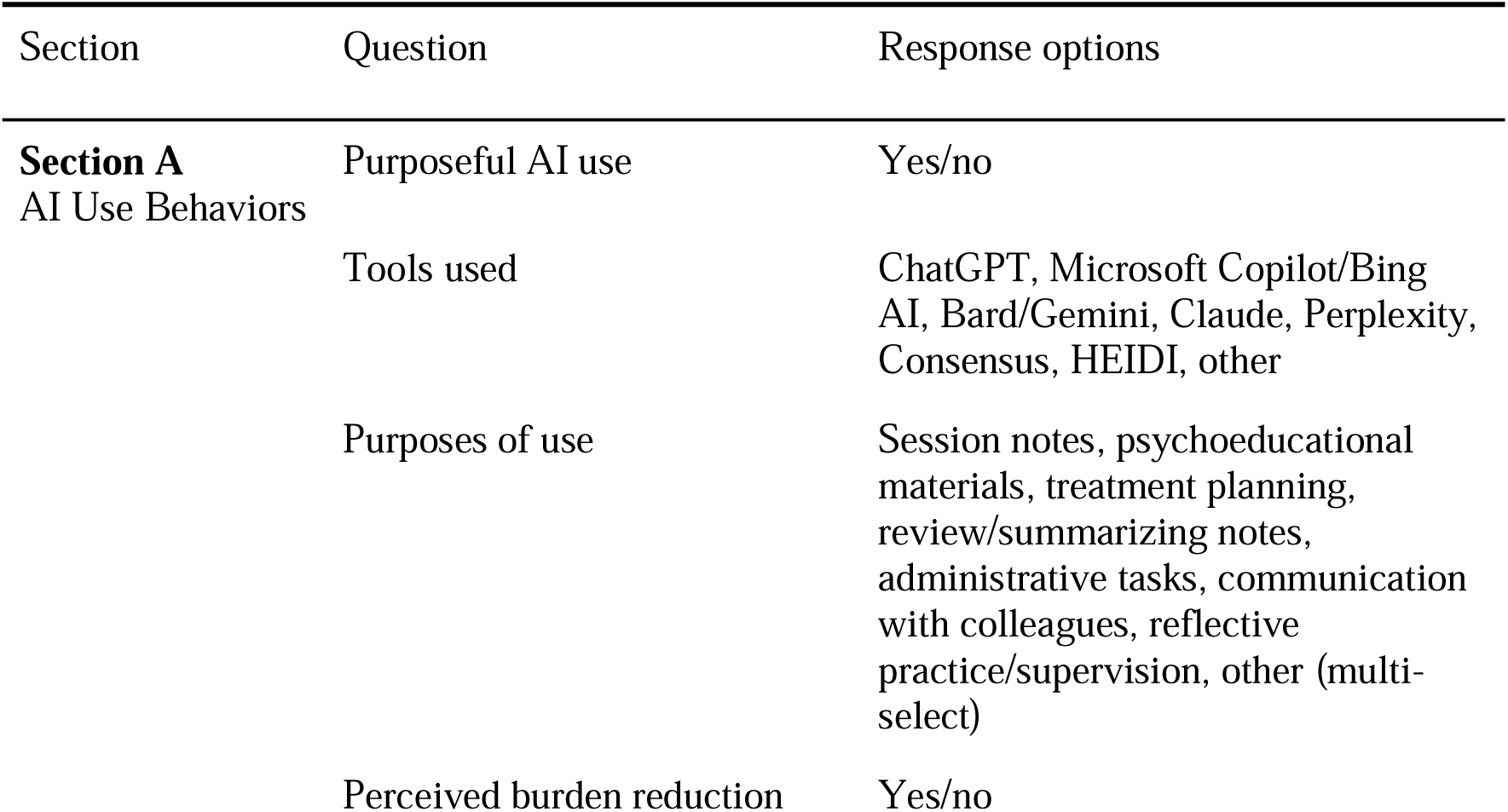
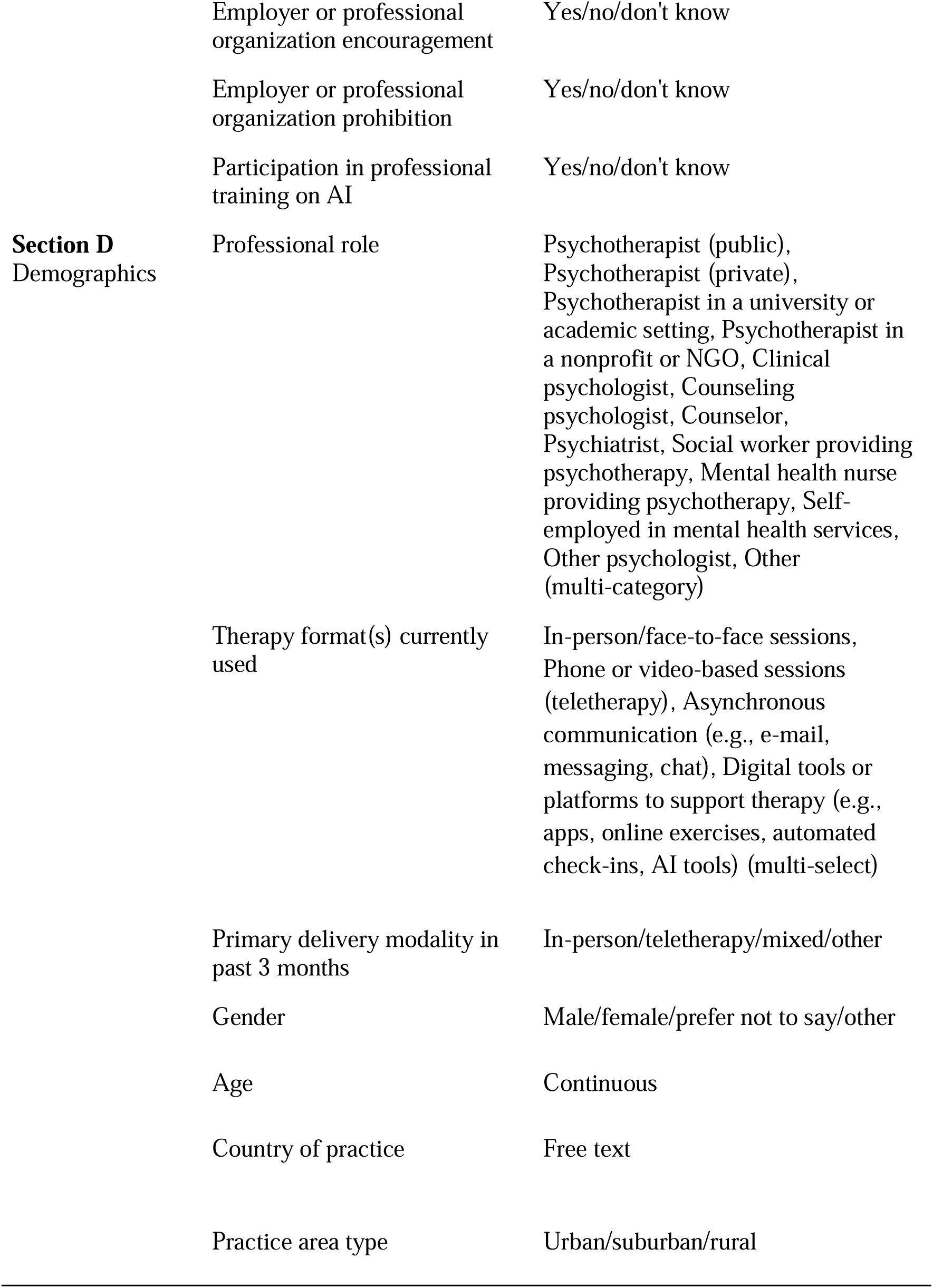
Reported Sections of the Online Survey.

### Statistical Analysis

Data were exported from LimeSurvey. Descriptive statistics are reported as frequencies and percentages for categorical variables and means with standard deviations (SD) or medians with interquartile ranges (IQR) for continuous variables (age), depending on distribution normality assessed by the Shapiro-Wilk test.

Bivariable associations between AI use (yes/no) and candidate predictors (age, gender, professional role, primary delivery modality, practice setting, country income group [World Bank classification]) were examined using chi-square tests or Fisher’s exact tests for categorical variables and independent samples t-tests or Mann-Whitney U tests for continuous variables. Variables meeting an entry criterion of *p* < .10 in bivariable analyses were entered into a multivariable binary logistic regression model. Results are reported as adjusted odds ratios (aOR) with 95% confidence intervals (CI) and two-sided P values; significance threshold was set at *p* < .05. All analyses were conducted in SPSS Version 30.0 with listwise deletion for the regression model.

## Results

### Sample Characteristics

A total of 766 participants completed the survey (completion rate: 72.6% of those who opened the survey link (766/1055)). Respondents were located across 30 countries, predominantly from Germany (402/766, 52.5%) and the United States (87/766, 11.4%). Other reported countries were: Argentina, Australia, Austria, Belgium, Brazil, Bulgaria, Canada, Chile, Colombia, Denmark, Finland, France, Greece, Hungary, Ireland, Israel, Italy, Japan, Netherlands, New Zealand, Portugal, Romania, Slovenia, Spain, Sweden, Switzerland, UK, and Ukraine. A full list of countries and detailed demographic and professional characteristics of the sample are presented in Table 2.

**Table 2.** Demographic and Professional Characteristics of the Sample (N = 766)

| Characteristic | n | % (95% CI) |
| --- | --- | --- |
| ***Gender*** |  |  |
| Female | 510 | 66.6 |
| Male | 235 | 30.7 |
| Other / Prefer not to say | 16 | 2.1 |
| Age, years — mean (SD) / median (IQR) / range | 47.81 (14.03) / 46.50 (21) / 22-89 | — |
| **Country** |  |  |
| Germany | 402 | 52.5 |
| US | 87 | 11.4 |
| Ireland | 53 | 6.9 |
| Switzerland | 49 | 6.4 |
| UK | 33 | 4.3 |
| Canada | 25 | 3.3 |
| Greece | 23 | 3.0 |
| Austria | 14 | 1.8 |
| Denmark | 12 | 1.6 |
| Romania | 11 | 1.4 |
| Belgium | 9 | 1.2 |
| Argentina | 5 | 0.7 |
| Australia | 5 | 0.7 |
| Portugal | 5 | 0.7 |
| France | 3 | 0.4 |
| Israel | 3 | 0.4 |
| Japan | 3 | 0.4 |
| Spain | 3 | 0.4 |
| Ukraine | 3 | 0.4 |
| Brazil | 2 | 0.3 |
| Chile | 2 | 0.3 |
| Italy | 2 | 0.3 |
| Sweden | 2 | 0.3 |
| Bulgaria | 1 | 0.1 |
| Colombia | 1 | 0.1 |
| Finland | 1 | 0.1 |
| Hungary | 1 | 0.1 |
| Netherlands | 1 | 0.1 |
| New Zealand | 1 | 0.1 |
| Slovenia | 1 | 0.1 |
| Missing | 4 | 0.5 |
| ***Professional Role*** |  |  |
| Psychotherapist (public) | 194 | 25.3 |
| Psychotherapist (private) | 187 | 24.4 |
| Clinical psychologist | 81 | 10.6 |
| Psychotherapist in a university or academic setting | 56 | 7.3 |
| Counsellor | 53 | 6.9 |
| Counselling psychologist | 27 | 3.5 |
| Psychiatrist | 24 | 3.1 |
| Psychotherapist in a nonprofit or NGO | 14 | 1.8 |
| Other (medical doctor, physician psychotherapist, self-employed, social worker, mental health nurse, or student/trainee, etc.) | 130 | 17.0 |
| ***Primary Delivery Modality (past 3 months)*** |  |  |
| Mostly in-person ($\geq 75\%$) | 619 | 80.8 |
| Mostly teletherapy ($\geq 75\%$) | 72 | 9.4 |
| Mixed ($\approx 40\text{--}60\%$ split) | 54 | 7.0 |
| Other | 21 | 2.7 |
| ***Practice Setting*** |  |  |
| Urban | 535 | 69.8 |
| Suburban | 134 | 17.5 |
| Rural | 97 | 12.7 |
| **World Bank Country Income Group** |  |  |
| High income | 751 | 98.0 |
| Upper middle income | 11 | 1.4 |
| Lower middle / low income | 0 | 0 |
| Missing | 4 | 0.5 |

*CI: confidence interval; IQR: interquartile range; SD: standard deviation. “Other” responses included mainly (1) professionals working in multiple roles or settings (e.g. private practice and academic, public healthcare and NGO), (2) professions not listed among the response options (e.g. occupational therapists, nurses, and physicians), and () trainees (e.g. individuals in psychotherapy, psychology, or counseling programs). The response options “Social worker providing psychotherapy”, “Mental health nurse providing psychotherapy”, “Self-employed in mental health services” were categorized as “Other” due to small cell sizes.*

### Prevalence and Tools Used

Overall, 418 of 766 (54.6%) of respondents reported having purposely used at least one generative AI tool in their psychotherapy practice. Among these users, ChatGPT was the most frequently used tool (354/418, 84.7%), followed by Google Bard/Gemini (74/418, 17.7%) and Microsoft Copilot/Bing AI (53/418, 12.7%). “Other” free-text responses primarily included VIA Health (4.3%), Mistral Le Chat (1.7%), and Grok (0.7%). Complete data on tool use frequency are presented in Table 3, and all free-text responses are provided in Table S1 in Multimedia Appendix 3.

**Table 3.** GenAI Tools Used by Mental Health Professionals Who Reported Use (n = 418)

| Tool | N | % of AI users |
| --- | --- | --- |
| ChatGPT (OpenAI) | 354 | 84.7 |
| Google Bard / Gemini | 74 | 17.7 |
| Microsoft Copilot / Bing AI | 53 | 12.7 |
| Perplexity | 43 | 10.3 |
| Claude (Anthropic) | 27 | 6.5 |
| VIA Health | 18 | 4.3 |
| HEIDI Health | 8 | 1.9 |
| Consensus | 8 | 1.9 |
| Other (specify) | 70 | 16.7 |

*Percentages sum to >100% as respondents could select multiple AI tools.*

### Purposes of GenAI Use

Among GenAI users, assisting with treatment planning was the most commonly endorsed purpose (175/418, 41.9%), followed by managing administrative tasks (173/418, 41.4%). Reviewing and summarizing prior client notes (17.2%) and enhancing communication with colleagues (72/418, 16.5%) were reported less frequently. Those who selected Other (94/418, 22%) described a variety of additional uses, where main ones were: text production and language, information search, professional reporting, clinical reasoning, and teaching/training. Full data are presented in Table 4, and sub-themes and frequencies for Themes for free-text responses are found in Table S2 in Multimedia Appendix 3.

**Table 4.** Purposes of Mental Health Professionals’ Use of Generative AI Tools (n = 418)

| Purpose | N | % of AI users |
| --- | --- | --- |
| Assisting with treatment planning | 175 | 41.9 |
| Managing administrative tasks | 173 | 41.4 |
| Generating psychoeducational materials for clients | 166 | 39.7 |
| Supporting reflective practice or supervision | 136 | 32.5 |
| Session note documentation | 128 | 30.6 |
| Reviewing and summarizing prior client notes | 72 | 17.2 |
| Enhancing communication with colleagues | 69 | 16.5 |
| Other | 94 | 22.5 |

### Perceived Workload Impact

Among those who had used GenAI tools, 82.8% (346/418) reported that these tools had reduced their work burden overall and 17.2% (72/418) felt AI tools had not reduced their burden.

### Institutional Context: Encouragement, Prohibition, and Training

Across all respondents, 18.1% (139/766) reported that their employer or professional organization had encouraged them to use GenAI tools in the past 12 months, while 12.7% (97/766) indicated they were unaware of any such encouragement. In contrast, 8.2% (63/766) reported that their organization had prohibited AI use. Formal professional training or workshops on GenAI were reported by only 18.3% (140/766) of respondents, while 81.1% (621/766) reported no such training. Data are presented in Figure 1.

**Figure 1.**
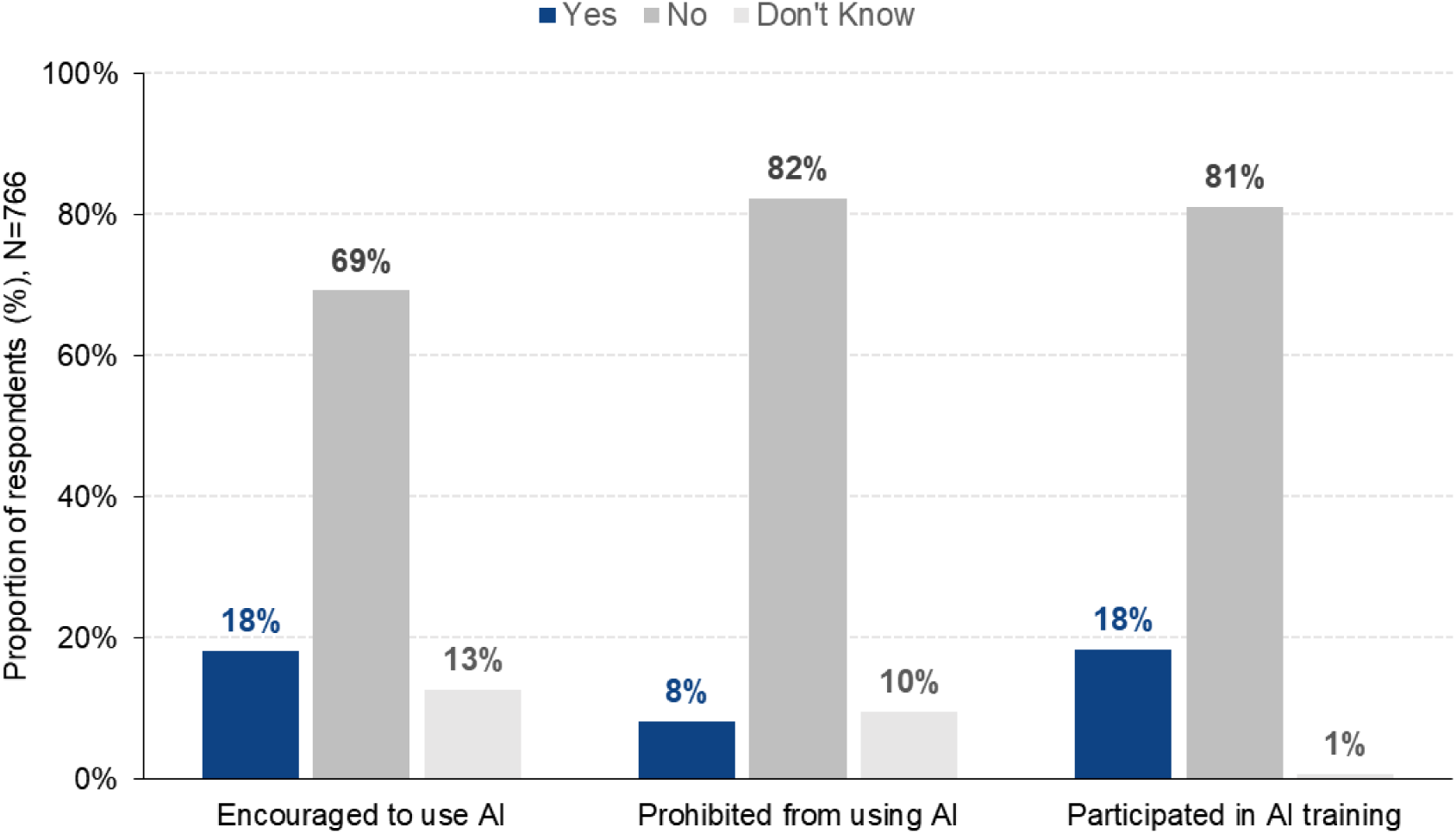
Proportion of respondents who have been encouraged to use GenAI, prohibited from using AI, or participated in AI-related training.

### Predictors of GenAI Use

In bivariable analysis, GenAI use was significantly associated with age (*p* < .001) and practice setting (*p* = .049). In the multivariable logistic regression model, independent predictors of AI use were: younger age (aOR=0.976, 95% CI 0.966–0.986, p<.001) and practice setting. Compared with those who reported they were rural practitioners, those in urban settings (aOR = 0.614, 95% CI 0.388–0.971, p=.037) and suburban settings (aOR = 0.537, 95% CI 0.311–0.930, p=.026) had significantly lower odds of GenAI use. In our sample, professional role, primary modality, gender, and country income group were not entered into the multivariable model as they did not meet the screening criterion of p < .10 in bivariable analyses. Full regression results are presented in Table 6.

**Table 6.** Multivariable Logistic Regression: Predictors of Purposeful GenAI Use Among Mental Health Professionals (N = 606)

| Variable | OR | 95% CI | P value | Adjusted OR | 95% CI | P value |
| --- | --- | --- | --- | --- | --- | --- |
| Age | 0.976 | 0.965-0.986 | <.001 | 0.976 | 0.966-0.986 | <.001 |
| Gender | 1.221 | 0.895-1.665 | .207 | — | — | — |
| Professional role: Private psychotherapist (ref) |  |  | .309 | — | — | — |
| Psychotherapist employed by a public healthcare system | 1.322 | 0.880-1.986 | .179 | — | — | — |
| Psychotherapist in a university or academic setting | 0.604 | 0.326-1.120 | .110 | — | — | — |
| Psychotherapist in a nonprofit or NGO | 0.851 | 0.287-2.523 | .772 | — | — | — |
| Counsellor | 0.820 | 0.445-1.510 | .524 | — | — | — |
| Clinical psychologist | 1.119 | 0.662-1.892 | .675 | — | — | — |
| Counselling psychologist | 1.448 | 0.630-3.327 | .384 | — | — | — |
| Psychiatrist incorporating psychotherapy in practice | 1.192 | 0.504-2.820 | .689 | — | — | — |
| Setting: Rural (ref) |  |  | **.049** |  |  | .066 |
| Urban | 0.592 | 0.376-0.932 | **.023** | 0.614 | 0.388–0.971 | **.037** |
| Suburban | 0.531 | 0.310-0.911 | **.022** | 0.537 | 0.311–0.930 | **.026** |
| Modality: Mostly in-person ($\geq 75\%$ of sessions face-to-face) (ref) | | | .643 | — | — | — |
| Mostly teletherapy — live phone or video ($\geq 75\%$ remote) | 1.127 | 0.687-1.847 | .637 | — | — | — |
| About an even mix ($\approx 40\text{--}60\%$ split between in-person and teletherapy) | 0.805 | 0.461-1.404 | .444 | — | — | — |
| Country income group | 0.998 | 0.302-3.299 | .997 | — | — | — |

OR = odds ratio; aOR = adjusted odds ratio; CI = confidence interval; — = not entered into multivariable model (did not meet screening criterion of $p < .10$ ); Nagelkerke $R^2 = .047$ ; $n = 606$

## Discussion

### Principal Findings

This global survey of 766 mental health professionals characterizes the breadth and context of generative AI adoption in clinical psychotherapy practice across multiple countries. Our central finding, that approximately 54.6% of respondents have used GenAI tools in clinical practice, suggests that adoption is already substantial in this professionally and geographically diverse convenience sample, and likely growing. This rate is consistent with, and in some analyses exceeds GenAI-use estimates from concurrent surveys of physicians and clinical psychotherapists [2,4,15], suggesting that mental health professionals are not lagging behind other health disciplines in adoption of these tools. However, the findings show that, among mental health professionals, younger clinicians adopt the GenAI tools more readily.

The pattern of use is informative. Across tools adopted (predominantly ChatGPT, a general-purpose LLM) and the purposes endorsed (predominantly treatment planning, managing administrative tasks, psychoeducational content, and reflective practice), what emerges is a picture of AI integration. Clinicians may be turning to AI to reduce workplace burdens, but also for core clinical support. This raises important questions about how and whether these tools are effectively adopted, and how they might influence clinical accuracy and treatment outcomes [17–19]. Adoption also raises fundamental concerns about data privacy, particularly in countries with strict data protection frameworks [20,21].

The finding that a majority of AI users perceived a reduction in work burden aligns with emerging data from medicine which identifies ambient AI scribes as reducing burdens but not necessarily time spent on tasks such as documentation [22]. For mental health professionals, who face distinctive emotional labor, burnout risks, and extensive administrative demands, even modest efficiency gains in documentation may translate into meaningful restoration of time for direct clinical care or personal recovery [6]. Qualitative studies from psychotherapy settings further suggest that clinicians experience reduced emotional and cognitive burden, greater presence during sessions, and perceived improvements in documentation quality when using AI-supported documentation tools [23–25]

Against this backdrop of substantive adoption, institutional guidance is strikingly lacking. Only a small minority of respondents reported professional training on GenAI, and employer encouragement was inconsistent. This training gap is ethically consequential: without structured competency development, clinicians may be using tools whose limitations they are poorly positioned to detect or manage. Additionally, not aiding clinicians in the adoption of GenAI in their practice may be preventing them from the benefits these tools can bring to their well-being and reduced burnout risks as well as potential improvements in quality.

### Comparison with Existing Literature

Prior work on AI attitudes and behaviors in mental health has been limited in scope and geography [14]. A 2023 US-based survey found that ChatGPT adoption was high among surveyed psychiatrists [12]. Our survey is also consistent with American Psychiatric Association and American Psychological Association findings [4,15]. A European study of psychiatrists reported cautious attitudes toward AI in clinical decision-support, with concerns about liability and data security predominating [26] Our data extend these findings to a global, multi-professional sample and move beyond attitudes to characterize actual behaviors, tools, purposes, and institutional context. The dominance of ChatGPT in our sample, despite the availability of healthcare-specific tools such as Nuance DAX and HEIDI, suggests that cost, accessibility, familiarity, and institutional context may drive adoption, rather than clinical suitability. This is particularly concerning since the least expensive and most accessible tools are older versions of the models which have been shown to be more prone to hallucination (that is, the generation of content that is both compelling and fluent but factually inaccurate) [27,28]. Relatedly, emerging studies indicate that patients and the public are rapidly adopting GenAI tools for mental health and emotional support, in particular those with no health insurance, minorities, and younger people [29,30]. These studies suggest that ease of access of these tools, costs, waiting lists, and fear of stigmatization by health professionals are reasons for GenAI adoption.

### Implications for Policy and Practice

Our findings carry several implications for professional bodies, regulators, and training programs. First, competency frameworks for ethical and safe GenAI use in psychotherapy are urgently needed. The American Psychological Association, British Psychological Society, and equivalent bodies have begun to issue guidance, but formal curricula are sparse [31,32]. Second, the documented gap between informal adoption and institutional training or oversight represents a governance failure that needs to be addressed in licensing and continuing professional development frameworks. Third, the predominance of general-purpose, consumer LLMs over healthcare-specific tools in this sample suggests a need for better dissemination of information about purpose-built clinical AI options, including their regulatory status, data protection characteristics, and accuracy benchmarks.

### Limitations

There are several limitations with this study. First, this is a convenience sample recruited through professional networks, which likely overrepresents individuals with existing interest in digital health and AI, and therefore endorsing a higher prevalence of use relative to the broader global clinical workforce. While we specifically restricted the survey to professional networks, it was also not possible to verify employment status. A related concern is that many clinicians are already burned out and facing constrained time resources, and the additional burden of survey fatigue may have further biased the sample towards those with established interest in AI. Second, self-report data are also subject to social desirability, recall bias, and varying understandings of what constitutes GenAI tools. Third, our survey instrument was developed in English with a German translation. Owing to the resource constraints of the survey team, these linguistic limitations and language barriers limited participation from non-English-speaking and non-German-speaking professionals and skewed the geographic distribution of responses. Relatedly, owing to the snowball sample, and dependence on the team’s networks, the survey included participants from predominantly high-income, Western countries with few respondents based in low-income countries.

### Future Research

Future studies should examine the clinical outcomes associated with GenAI tool use in psychotherapy, particularly whether AI-assisted documentation affects note quality, session attentiveness, or therapeutic alliance ratings. Longitudinal designs are needed to track the trajectory of adoption and the impact of emerging professional training initiatives. A fruitful future research study would be to understand whether this is due to younger clinicians feeling more comfortable with the technology, or due to a lack of established efficient practices which come with more experience [33]. Qualitative exploration of the reasons practitioners endorse or avoid AI—including ethical reasoning, institutional norms, and practical barriers—would complement the quantitative profiling provided here. Specific investigation of GenAI use in low- and middle-income country contexts, where mental health workforce shortages are most acute, should be a priority. Finally, while out of the scope of the present study, but necessary to complement our findings, we strongly advocate for much greater research attention on client use of GenAI tools as emotional support tools, both as adjuncts to, and as replacements for, mental health clinicians.

## Conclusions

GenAI use among mental health professionals is already widespread in this global convenience sample, concentrated primarily in treatment planning, documentation and content generation tasks. It is notable that these uses are not only administrative but may influence the therapeutic content and process. Adoption substantially outpaces formal training, institutional guidance, and regulatory oversight. These findings are a call to action for professional bodies, health systems, and training programs to develop evidence-based competency standards, clear governance frameworks, and accessible educational resources that can support safe and ethically informed integration of AI into clinical mental health practice.

## Supporting information

Multimedia Appendix 1

Multimedia Appendix 2

Multimedia Appendix 3

## Acknowledgments

The authors thank all mental health professionals who participated in this survey. No specific funding was received for this study.

## Authors’ Contributions

CB and JCW conceived the study. CB, JCW, and JG contributed to survey design. CB, JCW, and JG contributed to ethical approvals. All authors oversaw survey administration and data collection. JH led the statistical analysis. CB drafted the manuscript; all authors reviewed and approved the final version.

## Data Availability

The preprint of this study is available on medRxiv and deidentified participant-level data are available upon request. The survey instrument is available as Multimedia Appendix 1.

## AI Statement

Artificial intelligence (AI) tools were used to assist with language editing and sentence refinement during manuscript preparation. The authors reviewed and revised all AI-assisted text and take full responsibility for the content of the manuscript.

**Multimedia Appendix 1: Survey instrument in English and German.**

**Multimedia Appendix 2: Checklist for Reporting Results of Internet E-Surveys (CHERRIES) checklist.**

**Multimedia Appendix 3: Free-text themes for "Tools Used" and "Purposes of Use".**

