## Supplementary material for "Generative Artificial Intelligence in Psychotherapy Practice: A Global Online Survey of Mental Health Professionals’ Adoption": Multimedia Appendix 1

### Multimedia Supplementary File 1: Survey Instrument

*This supplementary file contains the complete survey instrument (all sections). Section A and Section D are the items relevant to this paper and are reproduced below for reader reference.*

#### Section A: Use of Generative AI in Psychotherapy

- Q1a. Have you ever purposely used generative AI tools to assist you in any aspect of your psychotherapy practice? [Yes / No]
- Q1b. What tools have you used? [ChatGPT / Microsoft Copilot/Bing AI / Bard/Gemini / Claude / Med-PaLM / Nuance DAX / SUKI / HEIDI / Other] (conditional on Q1a = Yes)
- Q1c. For what purpose(s) have you used generative AI tools? [Session note documentation / Generating psychoeducational materials / Treatment planning / Reviewing/summarizing notes / Administrative tasks / Communication with colleagues / Reflective practice or supervision / Other] (conditional on Q1a = Yes)
- Q1d. In general, have these tools reduced your work burdens? [Yes / No] (conditional on Q1a = Yes)
- Q2a. In the last 12 months, has your employer or professional organization encouraged you to use generative AI tools in your work? [Yes / No / Don't know]
- Q2b. In the last 12 months, has your employer or professional organization prohibited you from using generative AI tools in your work? [Yes / No / Don't know]
- Q2c. In the last 12 months, have you participated in any professional training or workshops on the use of generative AI tools? [Yes / No / Don't know]

#### Section B: Opinions about Generative AI in Psychotherapy

- Q3. To what extent do you agree that generative AI tools will improve your work related to… [Strongly Disagree / Disagree / Somewhat Disagree / Somewhat Agree / Agree / Strongly Agree / Don't Know]
    - Client information gathering
    - Treatment planning
    - Clinical documentation
    - Enhancing therapeutic insights/reflections
    - Supporting clients outside of sessions (e.g., AI-driven journaling or chatbots)
    - Conveying empathy effectively in client communication
    - Understanding yourself to become more congruent with clients
    - Conveying unconditional positive regard effectively in client communication
    - Employing interventions in response to client markers, motivation, or requests
    - Communication with other healthcare professionals
- Q4. To what extent do you agree that the use of generative AI tools in psychotherapy will… [Strongly Disagree / Disagree / Somewhat Disagree / Somewhat Agree / Agree / Strongly Agree / Don't Know]
    - Increase client concerns around confidentiality
- Q5. As a result of generative AI, the meaningfulness of my job to me personally will… [Increase / Not change / Decrease]

- Q6. Please think about how your practice could be affected by generative AI. In your opinion, these tools will: [Decrease my risk of having legal action taken against me / Increase my risk of having legal action taken against me / Neither decrease nor increase my risk / Don't know]
- Q7. What percentage of your patients do you believe currently use generative AI tools for mental health-related purposes? [0% / 1-10% / 11-25% / 26-50% / 51-75% / 76-100% / Unsure]
- Q8. In general, do you believe generative AI tools are helpful or harmful for patients' mental health? [Strongly harmful / Somewhat harmful / Neither harmful nor helpful / Somewhat helpful / Strongly helpful / Unsure]
- Q9. To what extent do you agree with the following statement: "Generative AI will eventually replace most of the work currently done by human therapists." [Strongly Disagree / Disagree / Neither agree nor disagree / Agree / Strongly Agree]
- Q10. How soon do you think generative AI could replace most of the work done by therapists? [Within the next 5 years / Within 10 years / Within 20 years / More than 20 years from now / Unsure] (conditional on Q9 = Yes)

### Section C: Open-Ended Feedback

- F1. Please add any comments about this topic or the survey. [free-text]

### Section D: Demographic Information

- S1. Which of the following best describes your professional role? [multi-category: Private psychotherapist / Psychotherapist employed by a public healthcare system (e.g., NHS, Medicare, national health service) / Psychotherapist in a university or academic setting / Psychotherapist in a nonprofit or NGO / Counsellor / Clinical psychologist / Counselling psychologist / Psychiatrist incorporating psychotherapy in practice / Social worker providing psychotherapy / Mental health nurse providing psychotherapy / Self-employed in mental health services / Other psychologist (please specify below) / Other]
- S1a. Please specify type of psychologist: (conditional on S1 = Other psychologist)
- S2a. Which formats or tools do you currently use in your therapeutic practice? [multi-select: In-person / face-to-face sessions / Phone or video-based sessions (teletherapy) / Asynchronous communication (e.g., email, messaging, chat) / Digital tools or platforms to support therapy (e.g., apps, online exercises, automated check-ins, AI tools)]
- S2b. In the past 3 months, what has been your primary modality for delivering therapy? [Mostly in-person (≥75% of sessions face-to-face) / Mostly teletherapy — live phone or video (≥75% remote) / About an even mix (≈40–60% split between in-person and teletherapy) / Other]
- D1. What is your gender? [Male / Female / Prefer not to say / Other]
- D2. What is your age? [numeric]
- D3. Please specify the country where you work. [free-text]
- D4. Which of the following best describes the area where your practice is based? [Urban / Suburban / Rural]
