## Supplementary material for "Generative Artificial Intelligence in Psychotherapy Practice: A Global Online Survey of Mental Health Professionals’ Adoption": Multimedia Appendix 2

**Appendix 2.** Checklist for Reporting Results of Internet E-Surveys (CHERRIES)

| Item Category | Checklist item | Described in the manuscript | Cited from the manuscript |
|---|---|---|---|
| **Design** | Describe survey design | Yes | In **Methods > Study Design** |
| **IRB (Institutional Review Board) approval and informed consent process** | IRB approval | Yes | **Methods > Ethical considerations**<br>Ethical approval was obtained from the Institutional Review Board of the Faculty of Psychology, University of Basel (IRB number: 017-25-1) and Missouri State University's Institutional Review Board (IRB-FY2025-325). |
| | Informed consent | Yes | **Methods** > **Ethics Approval**<br>All participants provided informed consent prior to participation. |
| | Data protection | Yes | **Methods** > **Ethics Approval**<br>Data were collected using LimeSurvey, a secure online platform that employs encryption and anonymization to prevent survey responses from being linked to individual participants. All identifying information, including email addresses, was removed prior to the dataset being shared with the research team. The platform operates in compliance with the European Union General Data Protection Regulation (GDPR). |
| **Development and pre-testing** | Development and testing | Yes | **Methods > Survey Instrument**<br>The survey was developed iteratively by the lead members of the research team (CB, J C-W, and JG) with expertise in digital mental health, clinical psychology and psychotherapy, and survey methodology. Items were piloted with a convenience sample of three practicing clinicians from the US, UK, and Sweden before finalization. |
| **Recruitment process and description of the sample having access to the questionnaire** | Open survey versus closed survey | Yes | **Methods > Recruitment**<br>The survey link was disseminated directly and through snowball-sampling requests to professional contacts. Recruitment channels included professional associations and networks (psychology, psychiatry, counselling, and social work bodies), academic and clinical mailing lists. We deliberately avoided use of all social media platforms to administer the survey to avoid the risk of bot contamination. |

| | | | |
|---|---|---|---|
| | Contact mode | Yes | **Methods > Recruitment**<br>The survey link was disseminated directly and through snowball-sampling requests to professional contacts via e-mail. Recruitment channels included professional associations and networks (psychology, psychiatry, counselling, and social work bodies), academic and clinical mailing lists. |
| | Advertising the survey | Yes | **Methods > Recruitment**<br>The survey link was disseminated directly and through snowball-sampling requests to professional contacts via e-mail. Recruitment channels included professional associations and networks (psychology, psychiatry, counselling, and social work bodies), academic and clinical mailing lists. |
| **Survey administration** | Web/E-mail | Both | **Methods > Recruitment**<br>The survey link was disseminated directly and through snowball-sampling requests to professional contacts via e-mail. |
| | Context | Yes | **Methods > Recruitment**<br>Participants were recruited via a global convenience sampling strategy between 13 January 2026 and 30 March 2026 (approximately 7 weeks). The survey link was disseminated directly and through snowball-sampling requests to professional contacts via e-mail. Recruitment channels included professional associations and networks (psychology, psychiatry, counselling, and social work bodies), academic and clinical mailing lists. |
| | Mandatory/voluntary | Yes | **Methods > Ethics Approval**<br>All participants provided informed consent prior to participation. |
| | Incentives | Yes | **Methods > Ethics Approval**<br>Participation was voluntary, anonymous, and unpaid. |
| | Time/Date | Yes | **Methods > Recruitment**<br>Participants were recruited via a global convenience sampling strategy between 13 January 2026 and 30 March 2026 (approximately 7 weeks). |
| | Randomization of items or questionnaires | No | **--** |
| | Adaptive questioning | No | **--** |
| | Number of Items | Yes | **Methods > Survey Instrument** |

| | | | |
|---|---|---|---|
| | | | Section A — AI Use Behaviors (7 items)<br>Section D — Demographics (7 items) |
| | Number of screens (pages) | No | -- |
| | Completeness check | Yes | **Methods > Survey Instrument**<br>Participants were required to complete all closed-ended items for submission; a single open-ended free-text question was optional. |
| | Review step | No | |
| **Response rates** | Unique site visitor | No | Number not provided by LimeSurvey. |
| | View rate (Ratio of unique survey visitors/unique site visitors) | No | Number not provided by LimeSurvey. |
| | Participation rate (Ratio of unique visitors who agreed to participate/unique first survey page visitors) | Yes | **Results > Sample Characteristics**<br>The participation rate (unique visitors who agreed to participate / unique first survey page visitors) was 1,052 /1,116 (94.26%). |
| | Completion rate (Ratio of users who finished the survey/users who agreed to participate) | Yes | **Results > Sample Characteristics**<br>A total of 766 participants completed the survey (completion rate: 72.6% of those who opened the survey link). |
| **Preventing multiple entries from the same individual** | Cookies used | No | Not used as a means to prevent multiple entries. |
| | IP check | No | Not used as a means to prevent multiple entries. |
| | Log file analysis | No | Not used as a means to prevent multiple entries. |
| | Registration | No | Not used as a means to prevent multiple entries. |
| **Analysis** | Handling of incomplete questionnaires | No | -- |
| | Questionnaires submitted with an atypical timestamp | No | -- |
| | Statistical correction | No | -- |
