## Supplementary material for "Generative Artificial Intelligence in Psychotherapy Practice: A Global Online Survey of Mental Health Professionals’ Adoption": Multimedia Appendix 3

### Multimedia Supplementary File 3: Free-Text Themes for "Tools Used" and "Purposes of Use"

Table S1. Other generative AI tools mentioned in free-text responses and frequencies (question Q1b).

| Tool | n |
|---|---|
| Le Chat (Mistral) | 7 |
| Grok | 3 |
| PlaynVoice | 3 |
| Locally developed AI tool | 3 |
| Zoom AI Companion | 2 |
| Gamma | 2 |
| Otter AI | 2 |
| NotebookLM | 2 |
| Proton Lumo | 2 |
| Embedded genAI note-taking | 2 |
| Alpine AI | 1 |
| Aurora | 1 |
| BerichtBiber | 1 |
| Canva | 1 |
| Carelog | 1 |
| Terapeutbooking | 1 |
| DeepAI image generator | 1 |
| DeepL | 1 |
| Duck AI | 1 |
| Coachin | 1 |
| Elicit | 1 |
| Epic | 1 |
| Epikur KI | 1 |
| Grammarly | 1 |
| Klarify | 1 |
| Lindy | 1 |
| Logicc | 1 |
| Neuroflash | 1 |
| NOA (Jameda) | 1 |
| Open Evidence | 1 |
| Plaud | 1 |
| Scite.AI | 1 |

| | |
|---|---|
| SpeechAI | 1 |
| Teams AI | 1 |
| Upheal | 1 |
| Voilà | 1 |
| Wispr Flow | 1 |
| Non–AI-based applications (PVS System Tomedo, PDF, Dictation function in Microsoft Word) | 3 |
| “Different ones without login” | 1 |
| Don’t remember | 1 |

Table S2. Other purposes of AI tool use mentioned in free-text responses, examples, and frequencies (Q1c).

| Theme | Examples | n |
|---|---|---|
| Text production and language | Writing emails, blogs, reports, articles, etc | 27 |
| | Translation | |
| | Spelling, grammars | |
| | Formatting | |
| | Client communication | |
| Information search | Evidence for treatment | 17 |
| | Methods | |
| | Medical conditions | |
| | Technical terms and topics | |
| | Learning about client's cultural background | |
| | Clarification of legal issues | |
| Professional reporting | Case reports | 15 |
| | Protocol | |
| | Report (discharge) | |
| | Expert opinion | |
| Clinical reasoning | Explaining confidentiality | 13 |
| | Diagnostics | |
| | Case formulation | |
| | Test evaluation | |
| Teaching/training | Presentations | 7 |
| | Planning seminar structure | |
| | Assignments | |
| Material creation | Psychoeducational materials | 6 |
| | Script for explanation of confidentiality to clients | |
| | Creating treatment plans | |
| | Exercises | |
| | Graphics for client homework | |
| | Creating HIPAA-compliant forms | |
| Creativity | Inspiration | 6 |
| | Marketing | |
| | Social media posts | |
| | Ideas for therapy check-in and check-out tasks | |
| Administrative tasks | Organizing notes | 4 |
| | Creating school schedule | |
| | HIPAA form (legal requirements) | |
| Research support | Generating keyword terms/tags for research articles | 3 |
| | Reading suggestions and bibliography | |
| Technical support | Troubleshooting | 3 |
| | Structuring files | |
| Reflection | Reflection buddy | 2 |
| | Reflecting together with clients on their use of AI | |
